# LOW BIRTH WEIGHT AS A RISK FACTOR FOR SEVERE COVID-19 IN ADULTS

**DOI:** 10.1101/2020.09.14.20193920

**Authors:** Fàtima Crispi, Francesca Crovetto, Marta Larroya, Marta Camacho, Marta Tortajada, Oriol Sibila, Joan Ramon Badia, Marta López, Kilian Vellvé, Ferran Garcia, Antoni Trilla, Rosa Faner, Isabel Blanco, Roger Borràs, Alvar Agustí, Eduard Gratacós

**Author notes:** Fàtima Crispi and Francesca Crovetto contributed equally. Àlvar Agustí and Eduard Gratacós contributed equally. Correspondence and reprint requests: Fàtima Crispi. BCNatal - Barcelona Center for Maternal-Fetal and Neonatal Medicine (Hospital Clínic and Hospital Sant Joan de Deu), IDIBAPS, University of Barcelona, and Centre for Biomedical Research on Rare Diseases (CIBER-ER), Barcelona, Spain. Sabino de Arana 1, 08028 Barcelona, Spain.

## Abstract

The identification of factors predisposing to severe COVID-19 in young adults remains partially characterized. Low birth weight (LBW) alters cardiovascular and lung development and predisposes to adult disease. We hypothesized that LBW is a risk factor for severe COVID-19 in non-elderly subjects. We analyzed a prospective cohort of 397 patients (18-70y) with laboratory-confirmed SARS-CoV-2 infection attended in a tertiary hospital, where 15% required admission to Intensive Care Unit (ICU). Perinatal and current potentially predictive variables were obtained from all patients and LBW was defined as birth weight ≤2,500 g. Age (adjusted OR (aOR) 1.04 [1-1.07], P=0.012), male sex (aOR 3.39 [1.72-6.67], P<0.001), hypertension (aOR 3.37 [1.69-6.72], P=0.001), and LBW (aOR 3.61 [1.55-8.43], P=0.003) independently predicted admission to ICU. The area under the receiver-operating characteristics curve (AUC) of this model was 0.79 [95% CI, 0.74-0.85], with positive and negative predictive values of 29.1% and 97.6% respectively. Results were reproduced in an independent cohort, from a web-based survey in 1,822 subjects who self-reported laboratory-positive SARS-CoV-2 infection, where 46 patients (2.5%) needed ICU admission (AUC 0.74 [95% CI 0.68-0.81]). LBW seems to be an independent risk factor for severe COVID-19 in non-elderly adults and might improve the performance of risk stratification algorithms.

## INTRODUCTION

COVID-19 is a mild or asymptomatic condition in the majority of patients, but in up to 1-2% it may result in severe disease and death.^1,2^ Older age, male sex and coexisting conditions are the main risk factors described so far for severe COVID-19 disease.^3-8^ However, a small proportion of young and apparently healthy adults may eventually require critical care. There is a need for comprehensive models that identify factors associated to the risk of severe forms of COVID-19.^9^

The association between low birth weight (LBW) and adult health has long been recognized.^10-12^ LBW, defined as ≤2,500 g,^13-14^ can result from fetal growth restriction, prematurity or both.^13^ Fetal growth restriction has been associated with increased cardiovascular mortality,^10,15^ lower lung functional capacity^16-18^ and increased respiratory morbidity^18-19^ in adulthood. Likewise, prematurity has been described as a risk factor for suboptimal cardiovascular^20^ and lung^21^ development and a greater predisposition to heart failure^22^ and lung disease^23^ later in life. For studies in adults, birth weight is an accessible and robust surrogate for fetal growth restriction and preterm births, and a strong predictor of short and long-term morbidity.^24^

From the above observations, we hypothesized that LBW could increase the risk of developing severe illness in non-elderly adults with COVID-19. To test this hypothesis, we designed a prospective study in confirmed COVID-19 patients (18-70 years) admitted to our institution, a public, tertiary, referral, university hospital in Spain (development dataset) and validated the model in an independent cohort of self-reported laboratory-confirmed COVID-19 subjects recruited through a web-based survey (validation dataset).

## RESULTS

### Development dataset

Figure 1 (left panel) presents the CONSORT diagram of the testing cohort. Out of 516 potentially eligible patients (with laboratory-confirmed SARS-CoV-2 infection by real time polymerase chain reaction (RT-PCR) of nasopharyngeal swab samples) during the study period, the development cohort included 397 patients with available perinatal information (77%). Based on clinical assessment of severity, 98 patients (24.7%) followed home hospitalization care and 299 (75.3%) were hospitalized, 60 of whom (20%) were eventually taken care of in the Intensive Care Unit (ICU) (25 (42%) required mechanical ventilation and none died). Table 1 displays the characteristics of the 337 non-critically ill patients (home hospitalization (n=98) or in hospital (n=239)) with those treated in the ICU (n=60). The latter were older, more frequently males, had a higher body mass index (BMI) and a higher prevalence of hypertension. Of note, they were also born with LBW (18.3 *vs*. 9.5%, p=0.041) and suffered fetal growth restriction (25 *vs*. 14.8%, p=0.043) more often. Individuals born with LBW had a higher probability for ICU admission as compared with those with normal birth weight (Figure 2).

**Figure 1.**
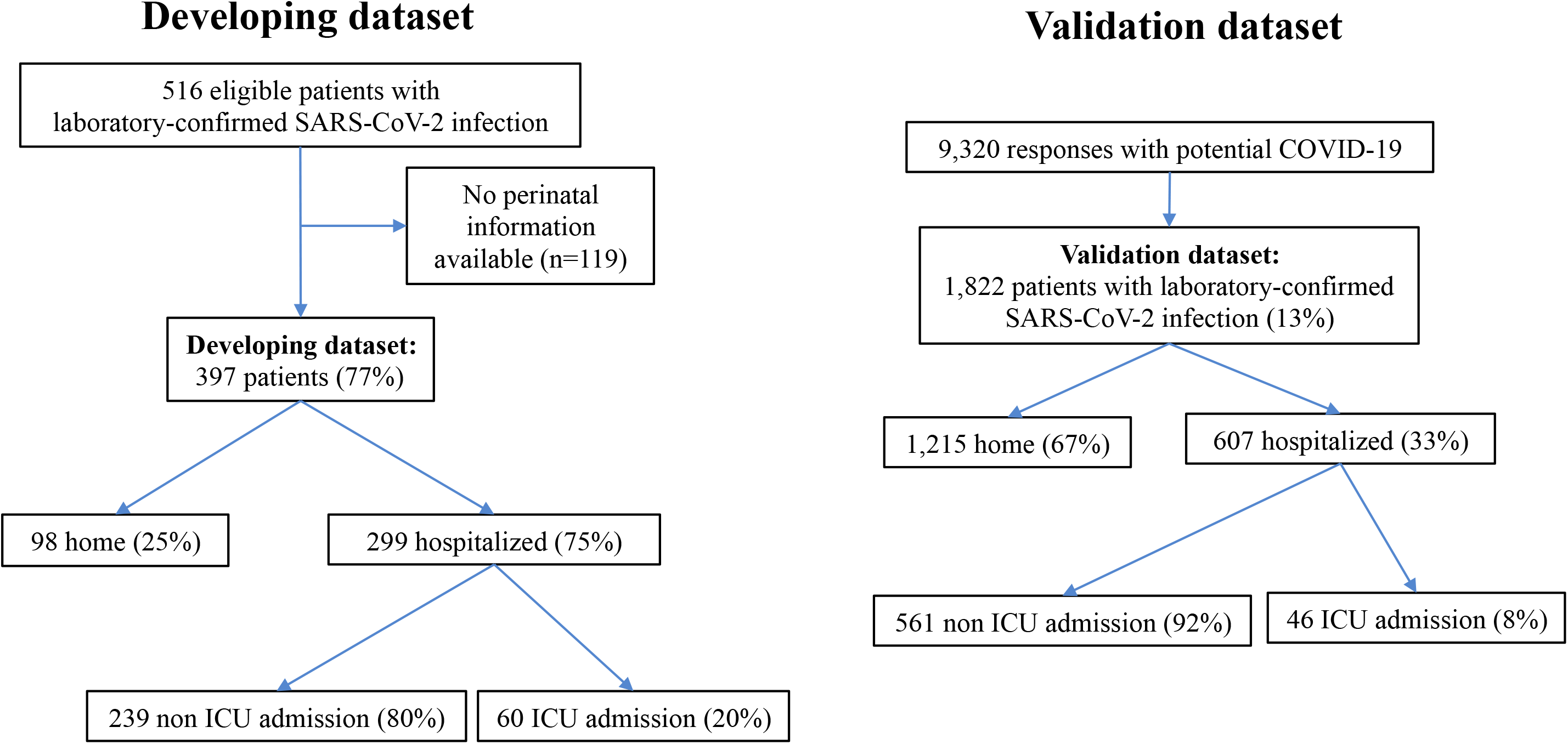
CONSORT diagram of the study.

**Figure 2.**
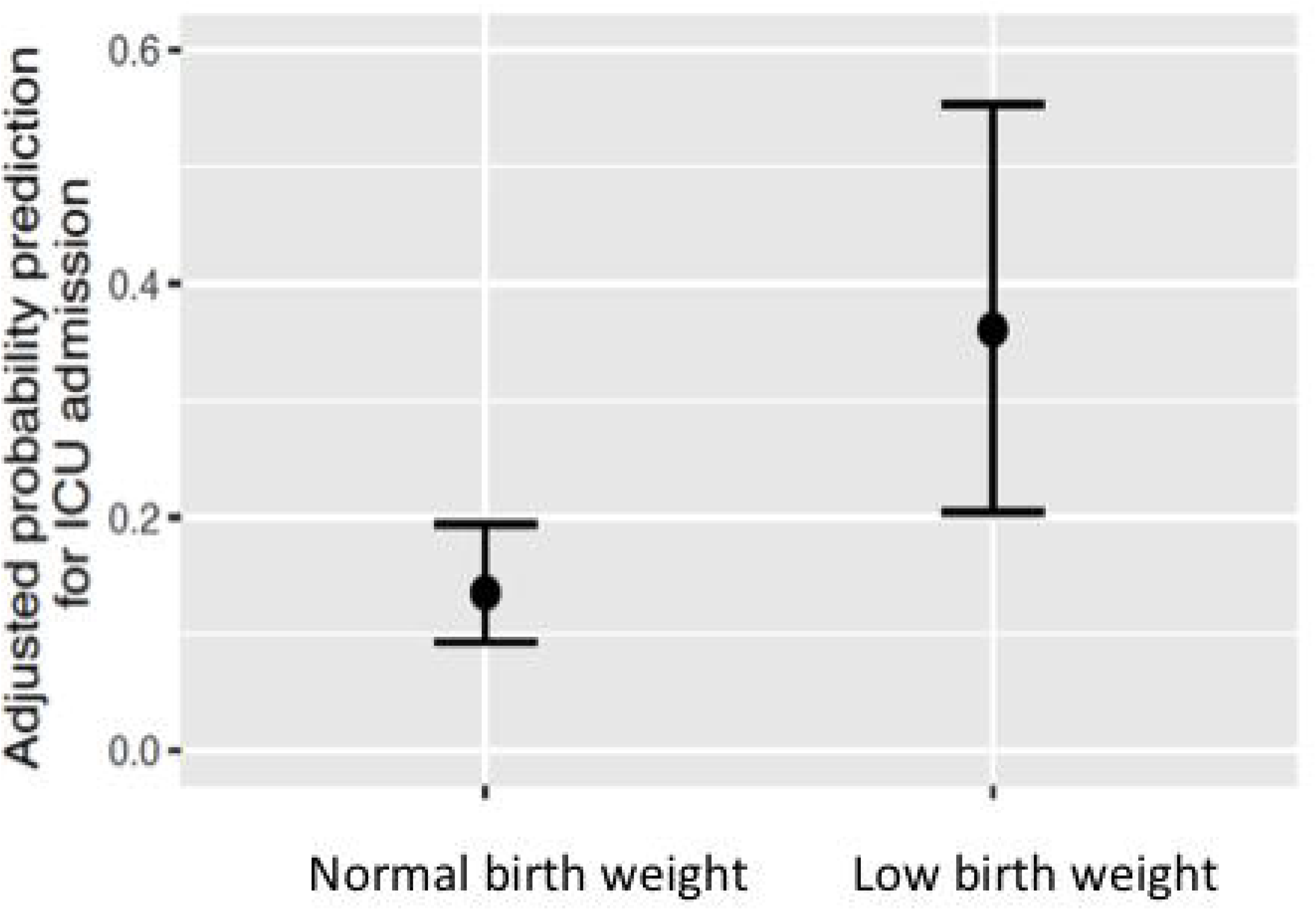
Adjusted probability prediction for ICU admission in normal *vs*. low birth weight individuals computed with multivariate logistic regression model. For further explanations, see text.

**Table 1.** Characteristics of participants in the development cohort by ICU admission.

|  | All population<br>(N=397) | No ICU<br>admission<br>(N=337) | ICU<br>admission<br>(N=60) | *P value\** |
| --- | --- | --- | --- | --- |
| *Demographic and clinical characteristics in adulthood* |  |  |  |  |
| Mean age (±SD) –yr | 47 ±12.2 | 46±12.2 | 53±10.1 | <0.001 |
| Female –no. (%) | 197 (49.6) | 183 (54.3) | 14 (23.3) | <0.001 |
| Mean body mass index (±SD) –Kg/m<sup>2</sup> | 26.9±5 | 26.6±5 | 28.6±4.8 | 0.005 |
| Current smoker –no. (%) | 27 (6.8) | 24 (7.1) | 3 (5) | 0.394 |
| Ex-smoker –no. (%) | 129 (32.5) | 104 (30.9) | 25 (41.7) | 0.069 |
| Coexisting conditions –no. (%) |  |  |  |  |
| Hypertension | 66 (16.6) | 40 (11.9) | 26 (43.3) | <0.001 |
| Cardiovascular disease | 16 (4) | 12 (3.6) | 4 (6.7) | 0.211 |
| Diabetes mellitus | 32 (8.1) | 26 (7.7) | 6 (10) | 0.350 |
| Obesity | 98 (24.7) | 84 (24.9) | 14 (23.3) | 0.467 |
| Dyslipidemia | 31 (7.8) | 23 (6.8) | 8 (13.3) | 0.077 |
| Chronic lung disease | 37 (9.3) | 33 (9.8) | 4 (6.7) | 0.312 |
| Chronic kidney disease | 20 (5) | 14 (4.2) | 6 (10) | 0.064 |
| Autoimmune disease | 12 (3) | 10 (3) | 2 (3.3) | 0.564 |
| Malignancy | 28 (7.1) | 22 (6.5) | 6 (10) | 0.235 |
| Thyroid disorders | 25 (6.3) | 21 (6.2) | 4 (6.7) | 0.540 |
| Other viral infections§ | 11 (2.8) | 8 (2.4) | 3 (5) | 0.223 |
| Psychiatric disorders | 23 (5.8) | 19 (5.6) | 4 (6.7) | 0.469 |
| Previous hospital admission within the last 12 months –no. (%) | 32 (8.1) | 27 (8) | 5 (8.3) | 0.548 |
| Drugs within last 15 days before Covid-19 diagnosis –no. (%) |  |  |  |  |
| NSAIDs | 142 (35.8) | 120 (35.6) | 22 (36.7) | 0.492 |
| ACE inhibitors | 27 (6.8) | 21 (6.2) | 6 (10) | 0.208 |
| Corticoids | 12 (3) | 11 (3.3) | 1 (1.7) | 0.436 |
| *Perinatal and childhood characteristics* |  |  |  |  |
| Mean birth weight (±SD) – g | 3302±666 | 3296±635 | 3335±826 | 0.728 |
| Low birth weight –no. (%) | 43 (10.8) | 32 (9.5) | 11 (18.3) | 0.041 |
| Fetal growth restriction –no. (%) | 65 (16.4) | 50 (14.8) | 15 (25) | 0.043 |
| Prematurity –no. (%) | 28 (7.1) | 23 (6.8) | 5 (8.3) | 0.420 |
| Childhood lung disease –no. (%) | 75 (18.9) | 67 (19.9) | 8 (13.3) | 0.155 |
| Asthma | 42 (10.6) | 39 (11.6) | 3 (5) | 0.229 |
| Bronchitis | 20 (5) | 16 (4.7) | 4 (6.7) | 0.126 |
| Other lung disease | 13 (3.3) | 12 (3.6) | 1 (1.7) | 0.579 |

ICU denotes Intensive Care Unit, NSAIDs non-steroidal anti-inflammatory drugs, and ACE Angiotensin-converting enzyme.
§ Other viral infections including HIV and/or hepatitis B or C.
\*P-value for the comparison of ICU admission vs. no ICU admission.

Table 2 displays crude and adjusted Odds Ratio (aOR) for ICU admission. In the multivariate model, age (aOR 1.04 [1-1.07], P=0.012), male sex (aOR 3.3 [1.72-6.67], P<0.001), hypertension (aOR 3.4 [1.69-6.72], P=0.001), and LBW (aOR 3.61 [1.55-8.43], P=0.003) remained independent predictors of ICU. As shown in Figure 3 (left), the area under the receiver-operating characteristics curve (AUC) for predicting ICU admission was 0.79 (95% CI, 0.74-0.85). The model had a sensitivity of 91.7%, specificity of 60.2%, positive predictive value (PPV) of 29.1% and negative predictive value (NPV) of 97.6% (Table 3).

**Table 2.** Odds Ratios for ICU admission for COVID-19 in the developing cohort.

|  | Univariate analysis |  | Multivariate analysis |  |  |
| --- | --- | --- | --- | --- | --- |
|  | OR (95% CI) | *p-value* | aOR (95% CI) | *p-value* | *Beta Coefficient* |
| Age, in 1-yr unit | 1.06 (1.03-1.09) | *<0.001* | 1.04 (1-1.07) | *0.012* | 0.03963 |
| Sex: male vs. female | 3.9 (2.07-7.37) | *<0.001* | 3.39 (1.72-6.67) | *<0.001* | 1.16734 |
| Body mass index, in 1 Kg/m<sup>2</sup> unit | 1.08 (1.02-1.13) | *0.005* |  |  |  |
| Smoker or ex-smoker: yes vs. no | 1.43 (0.82-2.48) | *0.206* |  |  |  |
| Hypertension: yes vs. no | 5.68 (3.09-10.43) | *<0.001* | 3.37 (1.69-6.72) | *0.001* | 1.23937 |
| Cardiovascular disease: yes vs. no | 1.93 (0.6-6.21) | *0.268* |  |  |  |
| Diabetes mellitus: yes vs. no | 1.33 (0.52-3.38) | *0.550* |  |  |  |
| Obesity: yes vs. no | 0.9 (0.5-1.75) | *0.792* |  |  |  |
| Chronic lung disease: yes vs. no | 0.8 (0.1-6.61) | *0.835* |  |  |  |
| Malignancy: yes vs. no | 1.59 (0.62-4.1) | *0.337* |  |  |  |
| Low birth weight: yes vs. no | 2.14 (1.01-4.52) | *0.046* | 3.61 (1.55-8.43) | *0.003* | 1.10971 |
| Fetal growth restriction: yes vs. no | 1.91 (0.99-3.69) | *0.053* |  |  |  |
| Prematurity: yes vs. no | 1.24 (0.45-3.4) | *0.675* |  |  |  |
| Childhood lung disease: yes vs. no | 0.62 (0.28-1.37) | *0.236* |  |  |  |
| Constant |  |  |  |  | -4.95673 |

ICU denotes Intensive Care Unit, OR Odds Ratio, aOR adjusted Odds Ratio, and CI confidence interval. To obtain ICU admission probability calculate $e^{\text{logit}}/(1+e^{\text{logit}})$ .

**Table 3.** Predictive accuracy of a multivariate model that includes age, sex, hypertension and low birth weight for ICU admission for COVID-19 in the developing cohort and validation dataset.

|  | **Patients admitted to ICU** | **Patients not admitted to ICU** | **Total patients** | **Predictive value** |
| --- | --- | --- | --- | --- |
| **Developing cohort** | (N=60) | (N=337) | (N=397) |  |
| Criteria positive§ | 55 True positive (29.1%) | 134 False positive (70.9%) | 189 | PPV, 29.1% |
| Criteria negative | 5 False negative (2.4%) | 203 True negative (97.6%) | 208 | NPV, 97.6% |
| Sensitivity | 91.7% |  |  |  |
| Specificity |  | 60.2% |  |  |
| **Validation dataset** | (N=46) | (N=1,780) | (N=1,826) |  |
| Criteria positive§ | 34 True positive (5.5%) | 582 False positive (94.5%) | 616 | PPV, 5.5% |
| Criteria negative | 12 False negative (1.0%) | 1198 True negative (99.0%) | 1210 | NPV, 99.0% |
| Sensitivity | 73.9% |  |  |  |
| Specificity |  | 67.3% |  |  |

ICU denotes Intensive Care Unit, PPV positive predictive value, and NPV negative predictive value.
§According to the logistic regression model, criteria positive is defined as a probability greater than 10.554%

**Figure 3.**
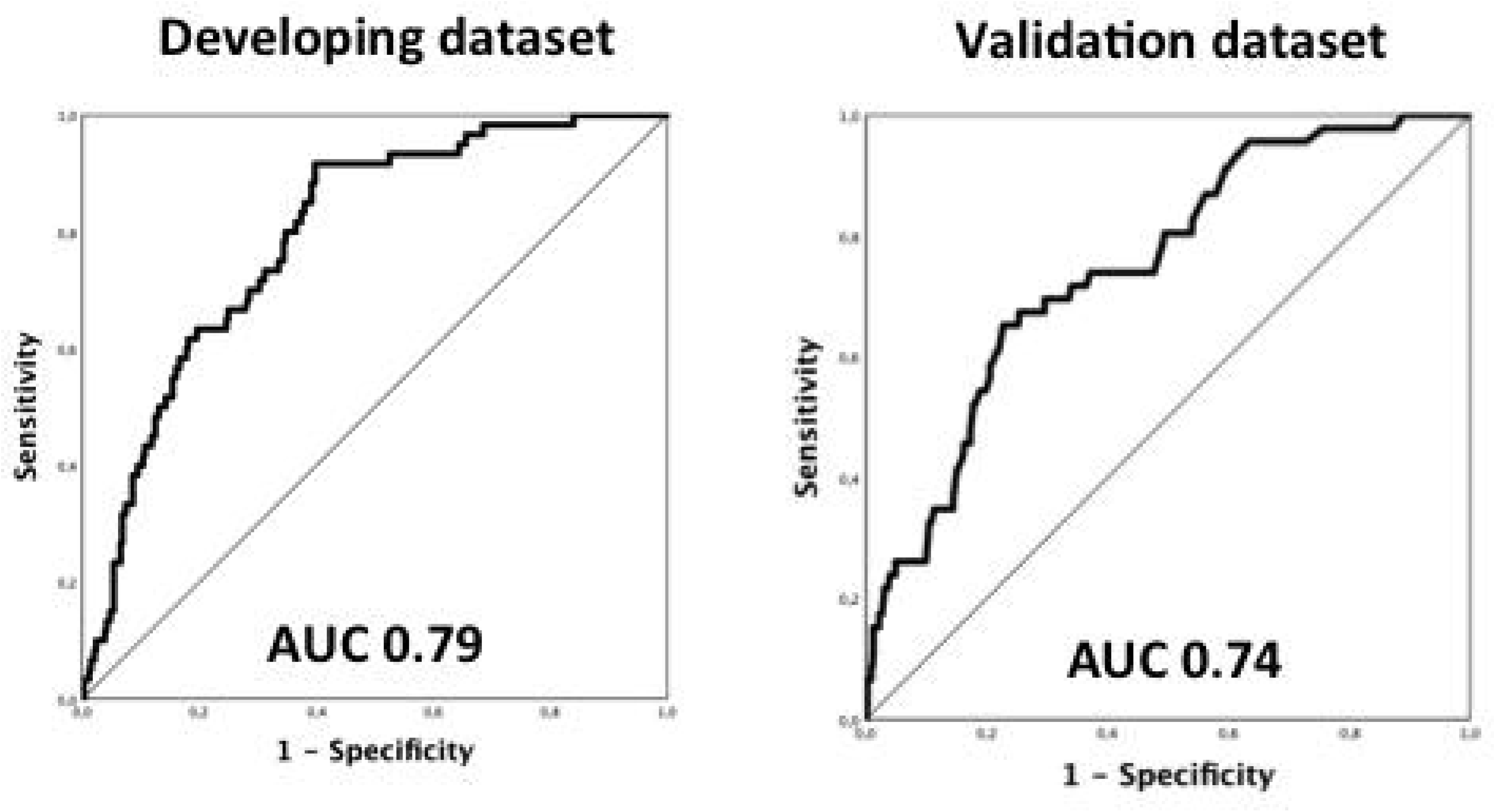
Receiver-operating characteristics curves for ICU admission in a multivariate model that includes age, sex, hypertension and low birth weight both in the development cohort (left) and validation dataset (right). For further explanations, see text.

### Validation dataset

Figure 1 (right panel) presents the CONSORT diagram of the validation dataset. We received 9,320 responses of subjects aged 18 to 70 years who referred symptoms suggestive of COVID-19. Among them, 1,822 self-reported COVID-19 confirmed by RT-PCR. A total of 1,215 of them (67%) reported mild symptoms and did not require hospital admission whereas 607 (33%) were hospitalized of whom 46 (8%) patients required ICU admission (30 of them (65% of ICU patients) were mechanically ventilated, and one male patient (2% of ICU patients) died at the age 68 years as reported later by her daughter.

Table 4 shows the characteristics of the 1,776 non-critically ill patients (treated at home (n=1,215) or in hospital (n=561)) with those treated in the ICU (n=46). Like we observed in the developing cohort, ICU patients in the validation dataset were older, more frequently males, had a higher BMI and a higher prevalence of hypertension. Importantly, again, they were born with LBW (19.6 *vs*. 7.3%, p=0.006) and suffered fetal growth restriction (26.1 *vs*. 12.4%, p=0.010) more often. In this validation dataset, the prevalence of prematurity was also higher in ICU patients (23.9 *vs*. 10.9%, p=0.011).

**Table 4.** Characteristics of COVID-19 patients in the validation dataset by ICU admission. ICU denotes Intensive Care Unit.

|  | All population<br>(N=1822) | No ICU<br>admission<br>(N=1776) | ICU<br>admission<br>(N=46) | *P value\** |
| --- | --- | --- | --- | --- |
| *Demographic and clinical characteristics in adulthood* |  |  |  |  |
| Mean age (±SD) –yr | 46 ± 11.5 | 46 ± 11.5 | 52 ± 9.1 | <0.001 |
| Female –no. (%) | 1255 (68.9) | 1239 (69.8) | 16 (34.8) | <0.001 |
| Mean body mass index (±SD) –Kg/m<sup>2</sup> | 25±4.9 | 24.9±4.8 | 28.9±6.3 | <0.001 |
| Current smoker –no. (%) | 132 (7.2) | 130 (7.3) | 2 (4.3) | 0.340 |
| Ex-smoker –no. (%) | 702 (38.5) | 682 (38.4) | 20 (43.5) | 0.290 |
| Coexisting conditions –no. (%) |  |  |  |  |
| Hypertension | 218 (12) | 208 (11.7) | 10 (21.7) | 0.041 |
| Cardiovascular disease | 55 (3) | 55 (3.1) | 0 (0) | 0.240 |
| Diabetes mellitus | 49 (2.7) | 46 (2.6) | 3 (6.5) | 0.124 |
| Obesity | 224 (12.3) | 210 (11.9) | 14 (30.4) | 0.001 |
| Dyslipidemia | 97 (5.3) | 95 (5.3) | 2 (4.3) | 0.552 |
| Chronic lung disease | 200 (11) | 196 (11) | 4 (8.7) | 0.419 |
| Chronic kidney disease | 17 (0.9) | 16 (0.9) | 1 (2.2) | 0.354 |
| Autoimmune disease | 93 (5.1) | 91 (5.1) | 2 (4.3) | 0.580 |
| Malignancy | 33 (1.8) | 32 (1.8) | 1 (2.2) | 0.573 |
| Thyroid disorders | 125 (6.9) | 120 (6.8) | 5 (10.9) | 0.204 |
| Psychiatric disorders | 61 (3.3) | 61 (3.4) | 0 (0) | 0.205 |
| *Perinatal and childhood characteristics* |  |  |  |  |
| Mean birth weight (±SD) –g | 3390 ± 606 | 3393 ± 595 | 3283 ± 935 | 0.432 |
| Low birth weight –no. (%) | 128 (7.6) | 129 (7.3) | 9 (19.6) | 0.006 |
| Fetal growth restriction –no. (%) | 233 (12.8) | 221 (12.4) | 12 (26.1) | 0.010 |
| Prematurity –no. (%) | 205 (11.3) | 194 (10.9) | 11 (23.9) | 0.011 |
| Childhood lung disease –no. (%) | 237 (13) | 233 (13.1) | 4 (8.7) | 0.826 |
| Asthma | 143 (7.8) | 141 (7.9) | 2 (4.3) | 0.309 |
| Bronchitis | 47 (2.6) | 46 (2.6) | 1 (2.2) | 0.340 |
| Other lung disease | 47 (2.6) | 46 (2.6) | 1 (2.2) | 0.666 |

ICU denotes Intensive Care Unit.
\*P-value for the comparison of ICU admission vs. no ICU admission.

The model obtained in development dataset was applied to the validation dataset, obtaining an AUC of 0.74 (95% CI 0.68-0.81) (Figure 2, right panel), with a sensitivity of 73.9%, specificity of 67.3%, PPV of 5.5% and NPV of 99% (Table 3).

## DISCUSSION

This study provides evidence that recording birth weight might improve the prognostic stratification of COVID-19 in non-elder patients. Most young patients present mild forms of COVID-19, but a small proportion might require admission to ICU for severe complications,^3-8^ which is clearly associated to non-obvious predisposing factors. Early interventions in COVID-19 have demonstrated to reduce mortality.^4,6^ Consequently, the identification of predisposing factors –particularly in a priori non high-risk subjects- might allow early therapeutic measures eventually preventing serious evolution to serious illness. In this study we evaluated an innovative approach by studying early life risk factors not usually taken into account in current clinical practices. Birth weight is one of the most universally recorded information for any given individual and self-recalled birth weight has demonstrated to be a reliable information, particularly in subjects born after the 1960s.^41^ If confirmed that LBW identifies high risk for complicated COVID-19, this should be included in initial assessment of non-elder infected subjects and would offer opportunities for early interventions to prevent complications.

### Previous studies

Despite the large number of studies on prognostic factors for severe COVID-19,^3-8,25-36^ to our knowledge no previous study has investigated the predictive role of early life events as a risk factor for severe COVID-19 in adulthood. Results confirmed our working hypothesis, which was aligned with a long-standing research line in this field.^11-12,18-19,37^ Besides, results confirmed previous studies showing a strong predictive value for severe COVID-19 of older age, male sex and coexisting conditions such as hypertension.^3-8,25-31^ The fact that we studied non-elderly adults (≤70 years) may have limited the identification of significant associations with other reported coexisting conditions such as chronic lung disease,^2,6-8,27^ diabetes,^2-5,7,8,25-26,30,32^ obesity^8,31,33^ or cancer.^3,34^ Current or previous smoking status^35^ and chronic treatment with ACE inhibitors^36^ were not associated with COVID-19 severity in our dataset.

### Interpretation of novel findings

Our results show that LBW is an independent risk factor for severe COVID-19 in adulthood. This finding is consistent with previous epidemiological and experimental studies supporting the developmental origin of adult diseases. Adverse i*n utero* environment induces permanent changes in the structure, function and metabolism of the developing fetal organs. Most developmental changes of early life persist in the long term which leads to a greater risk of disease in adulthood.^10,11^ It is suggested that fetal adaptation to perinatal events represents a ‘first hit’ leading to latent susceptibility, which combined with a ‘second hit’ later in life could increase the risk for adult diseases.^10,11^ This notion has been consistently demonstrated in experimental research,^40^ but evidence in humans is limited. The COVID-19 pandemic represents a unique opportunity to study the response of a significant number of individuals born LBW to a specific and well-defined stressor.

LBW has been consistently associated with increased adult cardiovascular mortality, hypertension, metabolic syndrome, diabetes and lung morbidity.^10,11,12,15-21^ LBW can be a result of being born too small –fetal growth restriction- and/or too early –prematurity-. Fetal growth restriction is caused by placental insufficiency leading to a sustained reduction in fetal oxygen and nutrient supply.^11^ This triggers an adaptive fetal response including cardiovascular remodeling,^12,36^ increased blood pressure,^12^ altered lipoprotein profile, lost of nephrons, and disturbed pulmonary alveolarization and vascular growth.^11^ In prematurity, key developmental stages have to take place *ex utero* in non-physiological conditions^39^ leading to cardiovascular hypertrophy and impaired lung development, insulin sensitivity and bone density.^20-22,39^

### Strengths and limitations

This study has some strengths and limitations that merit comment. Among the strengths, we prospectively collected information spanning the full COVID-19 clinical spectrum, from mild to hospitalized and ICU patients. Likewise, we validated our observations in an independent dataset. Finally, we included only non-elderly subjects (<70 years) to avoid the potential confounding effect of age-related comorbidities. The study sample size was too small to assess the predictive value of LBW across age ranges. We acknowledge that the evidence here presented should be validated in another prospective hospital cohort. We opted for an online survey to shorten validation time. We acknowledge also a potential selection bias since there were virtually no deaths in our study population. Firstly, mortality rate for COVID-19 was very low in our hospital (8%, 194/2,425) with most cases occurring in subjects >70 years-old. Secondly, we tried to contact all COVID-19 patients identified in the EMRs, but a few very severe cases were directly intubated and died preventing the interview for the study. Finally, we acknowledge the potential inaccuracy of the perinatal data obtained by interview or online survey. However, self-reported birth weight has demonstrated to be a good surrogate of adverse *in utero* environment, particularly in non-elderly subjects.^41^

### Conclusions

Low birth weight increases the risk of severe COVID-19 in non-elderly adults. This new information further supports the importance of early life events in adult diseases and should be considered in future risk stratification algorithms.

## METHODS

### Development dataset

#### Study design, Population and Ethics

Prospective observational cohort that included non-elderly adults (aged 18 to 70 years) consecutively attended at Hospital Clínic of Barcelona from March 25 to April 25, 2020 with laboratory-confirmed SARS-CoV-2 infection by real time polymerase chain reaction (RT-PCR) of nasopharyngeal swab samples. Sample size was determined by the time window of opportunity of the study. Criteria for hospital admission (COVID-19 pneumonia) and therapeutic management while in hospital followed the in-house protocols. The primary outcome of the study was admission to the ICU, which was determined by the attending physician on a patient by patient basis following standard clinical assessment criteria.^42^ Follow-up time was censored on May 25, 2020 so that each patient had at least 30 days of observation. The study was approved by the Ethical Committee of our institution (HCB/2020/0353) and informed consent was obtained from all patients.

#### Data collection

Cases were identified by daily review of hospital attendance logs in electronic medical records. Likewise, demographics, smoking exposure, coexisting conditions, treatment received during the last two weeks before hospitalization, need for ICU admission, complications or death during the clinical course, and therapeutic management interventions were obtained by reviewing electronic medical records. On the other hand, perinatal (birth weight and gestational age at delivery) and childhood (“asthma” or other respiratory disease in childhood) data were obtained by a face-to-face or telephone interview. Birth weight centiles were calculated adjusted by gender and gestational age at birth according to local standards.^43^ LBW was defined as birth weight equal or below 2,500g.^14^ Fetal growth restriction was defined as a birth weight below the 10^th^ centile for gestational age^44^ and preterm delivery as born before 37 weeks of gestation.^45^

### Validation dataset

#### Study design, setting and population

To validate externally the performance of the prognostic algorithm created by the development cohort, we collected independent data from self-selected volunteers who declared laboratory-confirmed SARS-CoV-2 infection through an anonymous multilingual (Spanish, Catalan, Italian, English and French) online survey (Limey Survey GmbH, Germany). The survey was disseminated via email and social media, and it was open and free for all subjects who self-reported to have laboratory confirmed COVID-19. Demographic information, coexisting conditions, perinatal and childhood data, COVID-19 related symptoms and need for hospitalization or admission to ICU were collected (voluntary sampling) using an anonymous web-based cross-sectional survey from April 1 to May 31, 2020 (LimeSurvey^®^).

### Statistical analysis

Results are presented as counts (percentage) or mean (SD) as appropriate. Variables with p<0.05 on univariate analyses were entered in the multivariate logistic regression analysis to determine independent risk factors for ICU admission. A forward stepwise selection algorithm was applied to select the final model in the development dataset. Odds ratio and 95% confidence interval [95%CI] were calculated. Hosmer and Lemeshow test were used to assess the goodness of fit of the final model.^46^ Analysis of the Receiver Operating Curve (ROC) was used to evaluate the predictive performance of the model in the development datasets and the optimal cut-off was computed using Youden criteria.^47^ The model determined in the development dataset was used to predict ICU admission in the validation dataset and we report the statistical parameters for development and validation. All p-values are 2-sided and considered statistically significant if <0.05. Data were analysed with SPSS v26 and R software version 3.6.2 (R project for statistical computing, Vienna, Austria).

## Data Availability

All data is available from the corresponding author at request.

## ACKNOWLEDGMENTS

Authors thank all participants in the study for their willingness to contribute to medical research as well as all the health workers who fought the COVID-19 pandemia.

